# School Teaching Posture Correlates with COVID-19 Disease Outcomes in Ohio

**DOI:** 10.1101/2021.07.16.21260464

**Authors:** Cheyenne Ehman, Yixuan Luo, Zi Yang, Ziyan Zhu, Sara Donovan, Annika J. Avery, Jiayi Wang, James Lawler, Rebecca Nugent, Valerie Ventura, Seema S. Lakdawala

**Affiliations:** Department of Statistics & Data Science, Carnegie Mellon University, Pittsburgh, PA, USA; Global Center for Health Security, University of Nebraska Medical Center, Omaha NE, USA; Department of Microbiology and Molecular Genetics, University of Pittsburgh School of Medicine, Pittsburgh PA, USA; Center for Vaccine Research, University of Pittsburgh School of Medicine, Pittsburgh PA, USA

## Abstract

At the start of the COVID-19 pandemic, most US K-12 schools shutdown and millions of students began remote learning. By September 2020, little guidance had been provided to school districts to inform fall teaching. This indecision led to a variety of teaching postures within a given state. In this report we examine Ohio school districts in-depth, to address whether on-premises teaching impacted COVID-19 disease outcomes in that community. We observed that counties with on-premises teaching had more cumulative deaths at the end of fall semester than counties with predominantly online teaching. To provide a measure of disease progression, we developed an observational disease model and examined multiple possible confounders, such as population size, mobility, and demographics. Examination of micropolitan counties revealed that the progression of COVID-19 disease was faster during the fall semester in counties with predominantly on-premises teaching. The relationship between increased disease prevalence in counties with on-premises teaching was not related to deaths at the start of the fall semester, population size, or the mobility within that county. This research addresses the critical question whether on-premises schooling can impact the spread of epidemic and pandemic viruses and will help inform future public policy decisions on school openings.

## Main text

The impact of school closures on community spread and mortality of COVID-19 is largely unknown. During the COVID-19 pandemic school posture has been thought to have a low impact on COVID-19 public health outcomes based on analysis from large-scale epidemiological studies^1,2^ and the low incidence of SARS-CoV-2 in school age children^3^. However, children are known to act as a vector for influenza virus transmission^4^ and given the lack of testing in asymptomatic individuals it is feasible that infections in kids have gone undiagnosed.

Each school district across the country chose their own teaching method with input from state governments. But the lack of federal or state mandates created scenarios where school districts within a single state had different teaching postures, either online (or virtual), hybrid format, or completely on-premises. The initial teaching mode for each school district across the US was captured by MCH Strategic Data (www.mchdata.com/covid19/schoolclosings), with verification occurring on a rolling basis throughout the Fall 2020 academic semester. Supplemental Fig 1A highlights the heterogeneity of teaching methods across the US school districts. The diversity of teaching methods for each state can be leveraged to examine the relationship between teaching posture, on-premises, hybrid, or online and its impact on COVID-19 deaths within the community.

In this study, we analyzed the impact of school teaching posture on COVID-19 deaths in Ohio. Ohio was selected for its small size with uniform geography and climate as well as two other important reasons. First, multiple public health interventions including mask orders, restaurant/bar restrictions, and gathering ban sizes were declared by the Governor and implemented statewide (Supplemental Fig 2, www.PhightCOVID.org). The statewide implementation of interventions allow for direct comparison between counties without the confounder of public-health mandate disparity. Second, Ohio school districts had diverse teaching postures (Supplemental Fig 1B), which allows for comparison between different teaching methods. The vast majority of counties within Ohio have similar population densities, but different COVID-19 death incidences based on population size (Supplemental Fig 3). Each county within Ohio has multiple school districts within it, although not all school districts within a county had the same teaching posture (Fig 1). Each county was categorized based on the largest proportion of students in a specific teaching mode. This strategy led to 16 ‘on-premises’, 11 ‘online only’, and 59 ‘hybrid model’ counties (Fig 1D).

**Figure 1.**
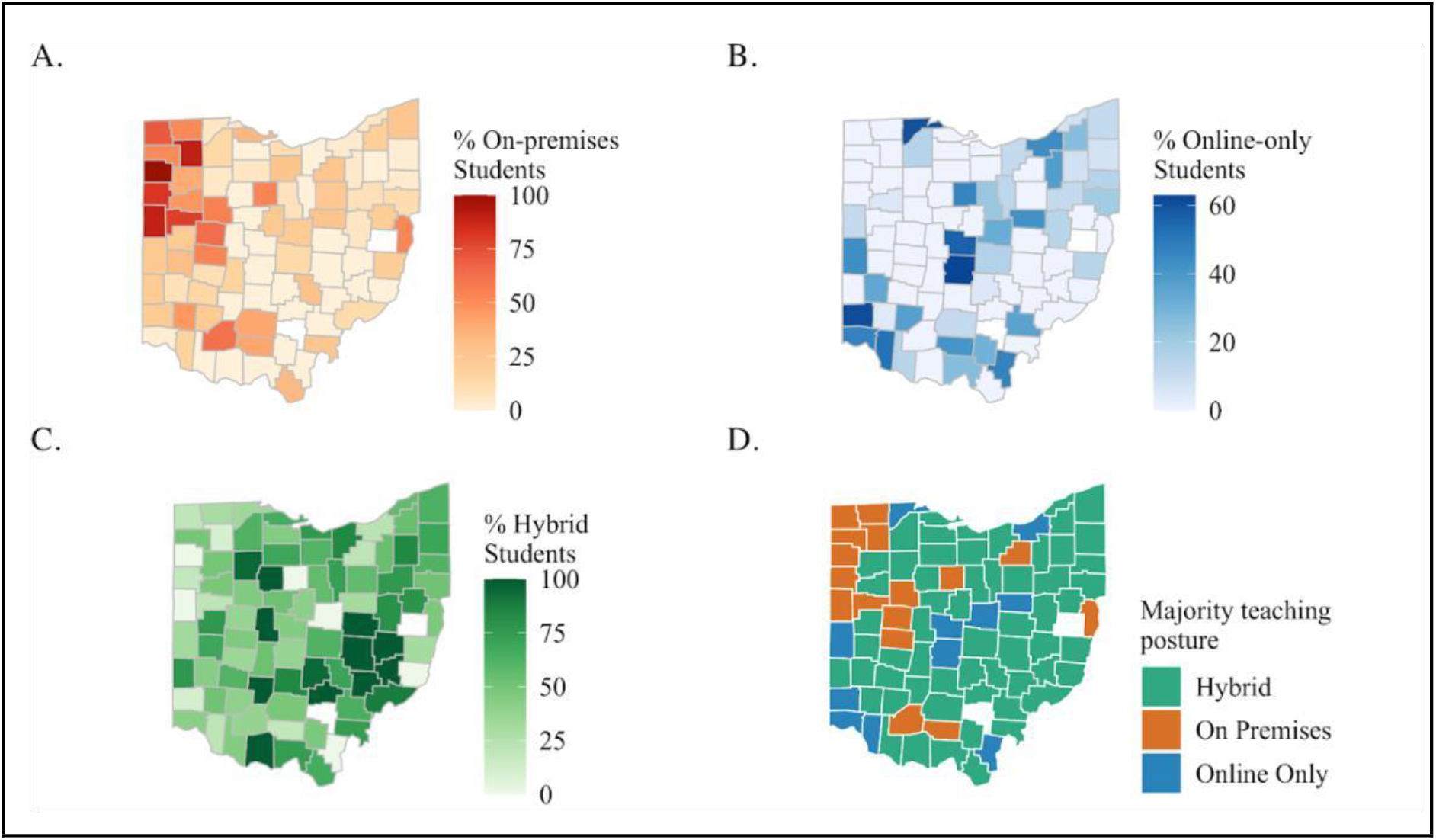
Distribution of teaching posture in Ohio counties. The percent of students within a given Ohio county that are A) On-premises, B) Online, or C) Hybrid. D) Each county was categorized for its teaching posture, based on the largest proportion of students in a specific teaching mode. Counties categorized as ‘on-premises’ are in orange (n=16), ‘online’ are in blue (n=11), and ‘hybrid’ (n=59) in green.

The cumulative COVID-19 deaths per 1,000 individuals was examined based on the primary teaching method in that county (Fig 2). The deaths in counties where the primary teaching mode was online or hybrid were higher in the summer months than those with on-premises teaching (Fig 2A). The levels of COVID-19 deaths in the community likely influenced the teaching method decision for the coming academic year within a given county. However, during the course of the fall term and after, there is a larger increase in the incidence of COVID-19 death in counties with predominate on-premises teaching compared to online teaching (Fig 2A). A comparison of all counties based on teaching posture and the cumulative deaths per 1,000 individuals throughout the fall semester indicate a significant difference between on-premises teaching and hybrid/online teaching (Fig 2C). This is a striking observation, but may be misleading given potential confounders within each county, such as the differences in deaths prior to the start of school that may have influenced not only the teaching method implemented but also the subsequent trajectory of COVID cases. To further explore the relationship between teaching posture and the counties’ disease state, we developed a generative model to better characterize the instantaneous progression of the disease.

**Figure 2.**
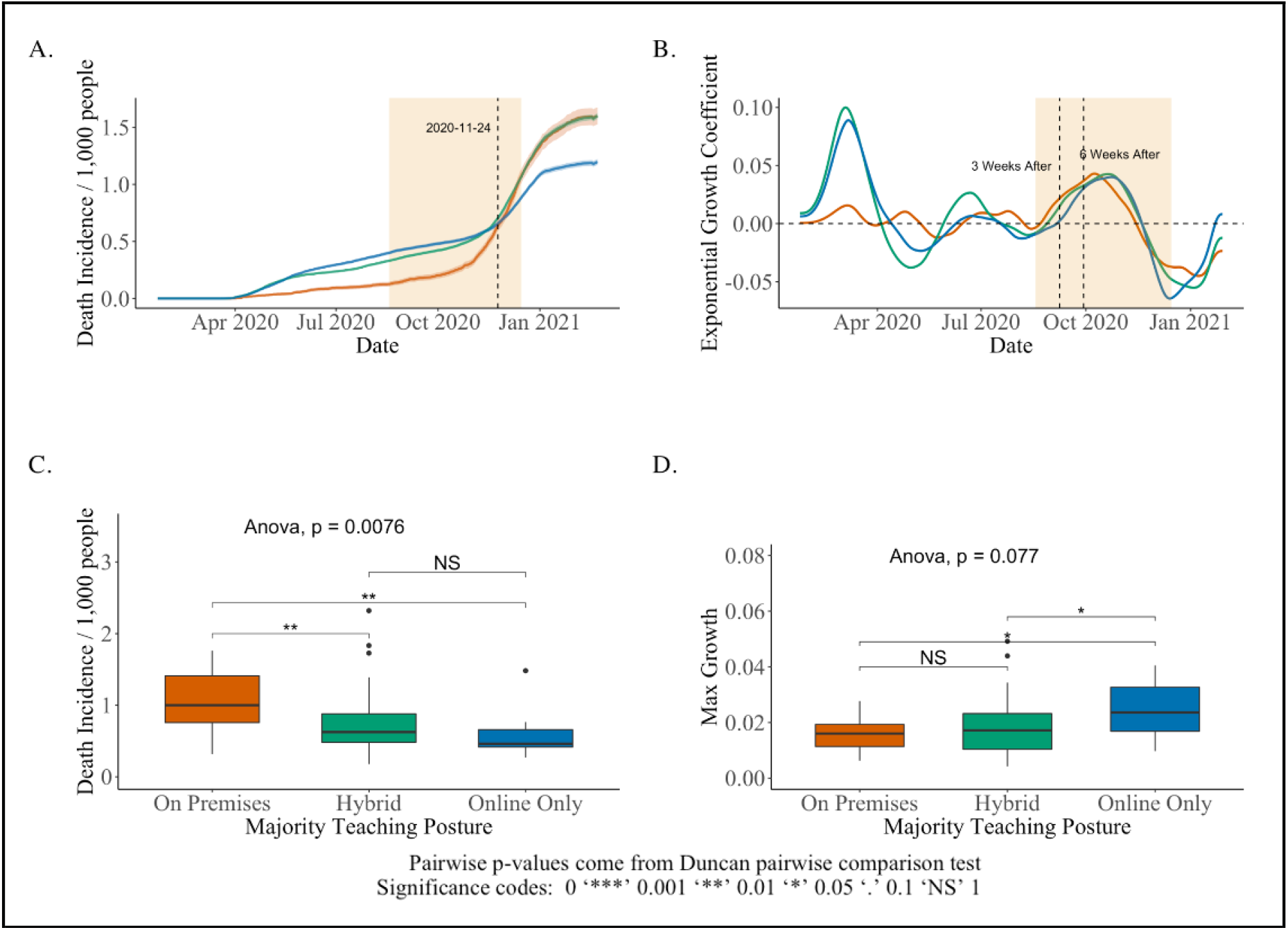
Incidence of death in Ohio counties based on majority teaching posture. A) COVID-19 deaths normalized to the population of all counties with a specific teaching method (on-premises, hybrid or online) are displayed from March 2020-Feb 2021. The lighter color shading surrounding each line indicates the 95% Gaussian pointwise confidence interval bands. A hatched line marks the Thanksgiving holiday. The fall school year term (from Aug 18 – Dec 15, 2020) is indicated by the yellow box. B) The change in COVID-19 disease prevalence based on an model fitted to observed deaths for all counties of a single teaching posture. Positive values indicate a growing pandemic and negative values indicate shrinking growth. The hatched line represents the start of school (Sept 8, 2020). The dashed black lines indicate 3 and 6 weeks into the fall semester. C) Boxplots of cumulative COVID-19 deaths per county during the fall semester based on predominant teaching posture. D) Boxplots of maximal exponential disease growth during the fall semester (highlighted yellow box in part B) for each county of a given teaching posture.

Our observational model describing the disease state within a given community is explained in detail in the Supplemental information. Briefly, observed daily deaths in each county are expressed as a function of the latent variable of new infections on day t (*I*_*t*_) and the equation *I*_*t*_ ≈ *exp*(*B*_*t*_) *I*_*t*−*1*_ models infections over time, which provides an easily interpretable value describing the growth of the disease. The growth factor is expressed as *exp*(*B*_*t*_) to ensure a positive value to fit the model to observed daily COVID-19 death data, where we can then obtain estimates of *B*_*t*_, herein referred to as ‘exponential growth coefficient’. A positive *B*_*t*_ implies that the number of new infections on day t exceeds the number of new infections on day t-1, indicating that the disease in that county is worsening. Conversely, a negative *B*_*t*_ suggests that the county’s disease state is improving. The exponential growth coefficients for all counties with a given teaching method were calculated and depicted via smoothed curves in Fig 2B. Curves above the y=0 line indicate time periods where *B*_*t*_ is positive, and thus *exp*(*B*_*t*_) > 1 and the disease state is worsening. When the curve dips below y=0, *B*_*t*_ is negative, *exp*(*B*_*t*_) <1, and the disease state is getting better.

Comparing the exponential growth coefficient for counties based on teaching method suggests that counties with online or hybrid teaching postures in the Fall of 2020 had expanding COVID-19 disease growth in March-April of 2020, (Fig 2B). However, at the start of the school year (Aug 2020) the pandemic in these counties was shrinking (Fig 2B, curves below y=0 dashed line) compared to those with on-premises teaching. In Oct of 2020, the three groups of counties were experiencing a similar growth in the pandemic. Counties with on-premises teaching had an increase in the growth of the pandemic earlier than the other counties, but the maximum growth in the three groups, all in mid-November, were similar (Fig 2B). Further exploration of the maximum disease growth (max of B1) for each county per teaching posture in the fall semester (Fig 2D) suggests that online counties had worse disease than on-premises counties. This observation is seemingly in contrast to analysis of cumulative deaths per 1,000 individuals (Fig 2C), but cumulative death measures all deaths from the entire fall semester where the max *B*_*t*_ represents the most severe growth of the disease at a single point in time. In addition, the disease state may also be influenced by confounders such as population size, mobility, and age of county residents such that comparison of the aggregated counties distorts the disease state present within individual counties. Thus exploring the variation in possible confounders and refining the analysis to counties with similar mobility, populations, or demographics are needed to better assess the effects of teaching posture on COVID-19 disease. Below we investigate the effects population density and mobility as potential confounders, but other covariates including county demographics and intervention adherence could impact the trajectory of COVID-19 deaths

To explore the role of mobility on COVID-19 disease during the Fall 2020 semester we utilized cell phone data from COVIDcast (https://delphi.cmu.edu/covidcast/). The COVIDcast data was used to calculate the proportion of cellphones within a given county that are away from their home zipcode for >6 hours a day, utilized as a metric for general mobility within a county. We observed that mobility in on-premises counties increased during the school term (yellow shaded box), likely due to the ability of parents to go to work places in person; whereas in online and hybrid counties, many parents may have to coordinate child care during the work day (Supplemental Fig 4). To incorporate increased mobility into the analysis of COVID-19 deaths and teaching method, the average mobility during the fall semester was plotted against the max disease growth coefficient during the fall semester per county (Fig 3A). Surprisingly, no positive correlation was observed between mobility and disease state, suggesting that the increased mobility, as measured by cell phone data, of the population may not be a strong driving force in disease outcomes. Rather mobility may be a consequence of the current disease state within a community.

**Figure 3.**
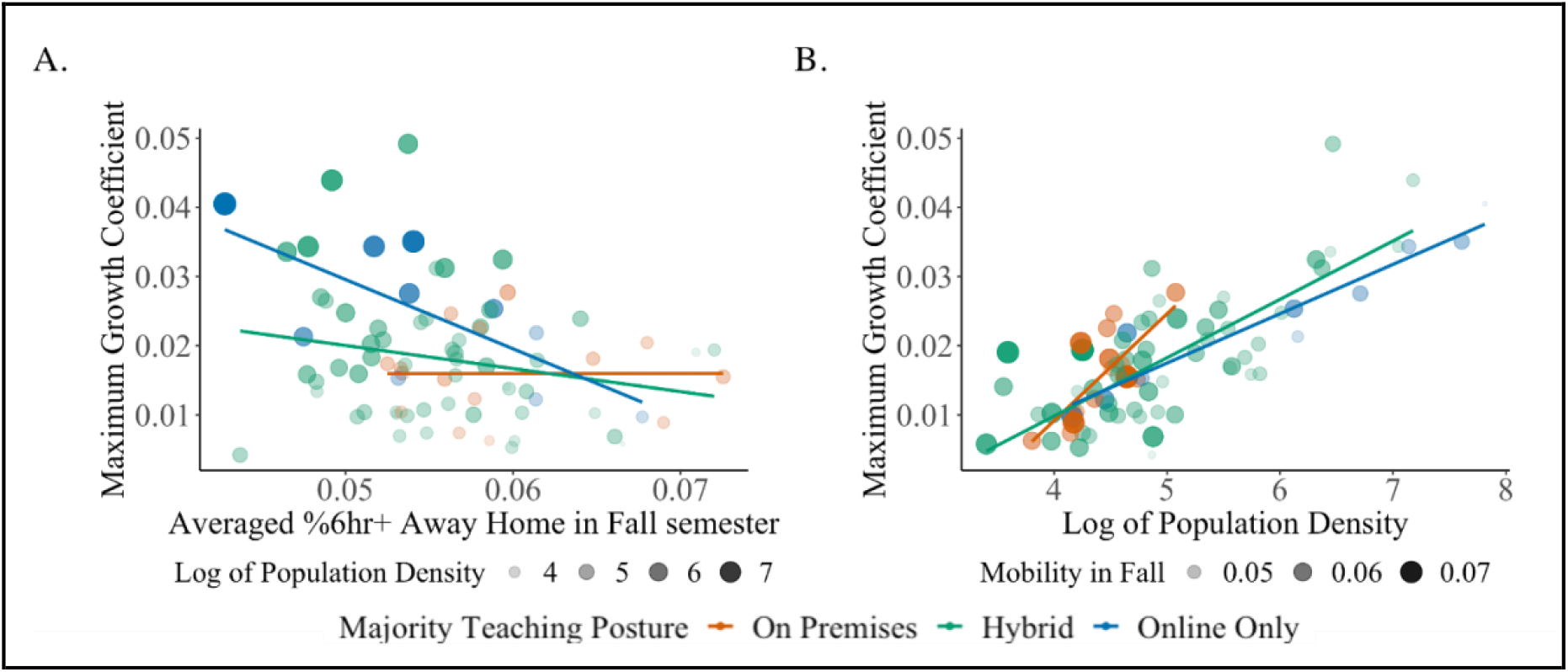
Maximum COVID-19 disease growth during the fall semester is related to population density, not mobility. The maximum growth coefficient during the fall semester from the exponential growth model for each county was calculated and compared to (A) the overall mobility of the county population or (B) the population density. Mobility data is based on cell phone data and aggregated to the proportion of county residents that spent 6+ hours away from their home during the Fall semester (COVIDcast data source). The population density was based on the census data (see methods for details on data sources).

The max disease growth coefficient during the Fall semester for each county was compared to the log population density for that county using linear regression for each teaching method group (Fig 3B). Consistent with other reports, we observed a correlation with population density and higher COVID-19 disease state^3,5^. Counties with higher population densities were primarily online or hybrid teaching methods. Looking at counties with log population density in the 4-5 range, the three teaching postures are represented and the maximum disease growth in on-premises teaching counties had a higher slope compared to the other teaching modes. This means that population density had a larger impact on COVID-19 deaths in the counties that chose on-premises teaching posture. However, this effect could be due to confounders rather than to teaching posture. For example, comparison of population age found that on-premises counties had more individuals >65, and based on the age disparity of COVID-19 deaths^6^ it is feasible that this would influence the increase in COVID-19 deaths (Supplemental Fig5). In addition, the proportion of uninsured individuals was higher in online counties compared to counties with primarily hybrid and on-premises, which could impact access to healthcare and/or behavior patterns (Supplemental Fig 5B).

In order to explore county demographics, we utilized the National Center for Health Statistics Urban-Rural classification scheme for counties based on the Office of Management and Budget delineation of metropolitan statistical areas and micropolitan statistical areas^7^. As expected, large and medium metropolitan areas were primarily online or hybrid (Supplemental Fig 6). Note that the large central metropolitan areas were largely online only while the suburbs to these large metropolitans (defined as large fringe metropolitans) tended to have more hybrid learning modes. Importantly, micropolitan counties, which are defined as urban areas with a population between 10,000-50,000, were distributed across teaching modes: on-premises (7), hybrid (23), and online (3) teaching counties (Fig 4A),which allows comparison of COVID-19 dynamics for the three teaching modes within a homogeneous group of counties. A similar analysis of the max disease growth coefficient restricted to micropolitan counties suggests that the epidemic got worse in on-premises counties (Fig 4B).

**Figure 4.**
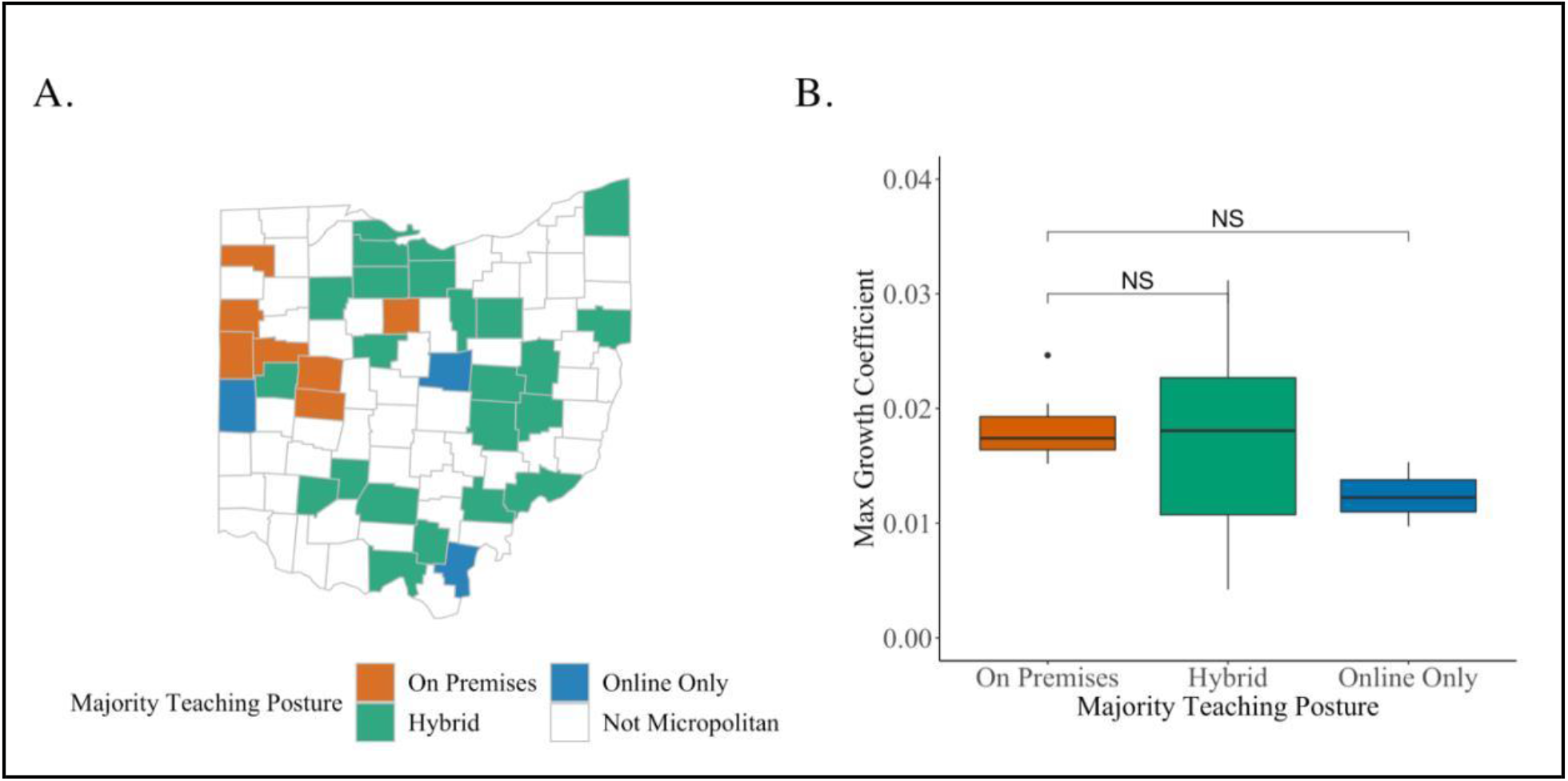
Analysis of COVID-19 disease in micropolitan counties in Ohio based on teaching method. A) Map of micropolitan counties in Ohio, color coded based on the teaching method. B) Box plots of maximum growth coefficient in micropolitan counties based on teaching mode.

Comparing the impact of teaching posture on max growth between all counties in Fig 2D and only micropolitan counties in Fig 4B reveals a striking difference. Fig 2D (max growth in all counties) suggests that online teaching counties had more COVID-19 disease during the Fall semester; however, in Fig 4B (only micropolitan counties) on-premises school posture is associated with higher disease spread. The apparent contradiction between Figs 2D and 4B is known as Simpson’s paradox^8^ and in this instance is due to the confounder of urban type. Urban type is associated with population density which may contribute to COVID-19 death incidence (higher incidence in more populous areas), adherence and interest in non-pharmaceutical interventions (NPI) compliance, and teaching posture. These two plots highlight the importance of accounting for confounders at multiple levels and strengthens the observations made by comparing micropolitan counties.

Another consideration, is that maximum disease growth during the fall semester occurred after the 6 week mark closer to Thanksgiving and may be influenced by other factors in addition to teaching posture. To narrow down the window on COVID-19 disease growth in the micropolitan counties, we examined the acceleration of COVID-19 disease at two different short time intervals at the beginning of the semester when covariates, other than school starting, are unlikely to have an impact. In this way, differences in disease growth observed can be attributed to school posture with reasonable confidence. First, we use the slope (or change in disease growth) over the first three weeks of the fall semester (week 0 – 3; first section of shaded box in Fig 2B) to explore differences in the disease state independent of teaching method. All micropolitan counties show similar behavior for change in disease for the first three weeks independent of teaching posture (Fig 5A). In contrast, the change in disease growth during the next three weeks (week 3-6) of the fall semester was consistently higher in counties with on-premises teaching compared to those with online or hybrid teaching (Fig 5B). That is, the acceleration of the disease was higher in counties with on-premises teaching compared to counties with hybrid and online-only teaching postures. This result is independent of mobility in each of these counties, as depicted by the size and color saturation in the data points in Fig 5A and B.

**Figure 5.**
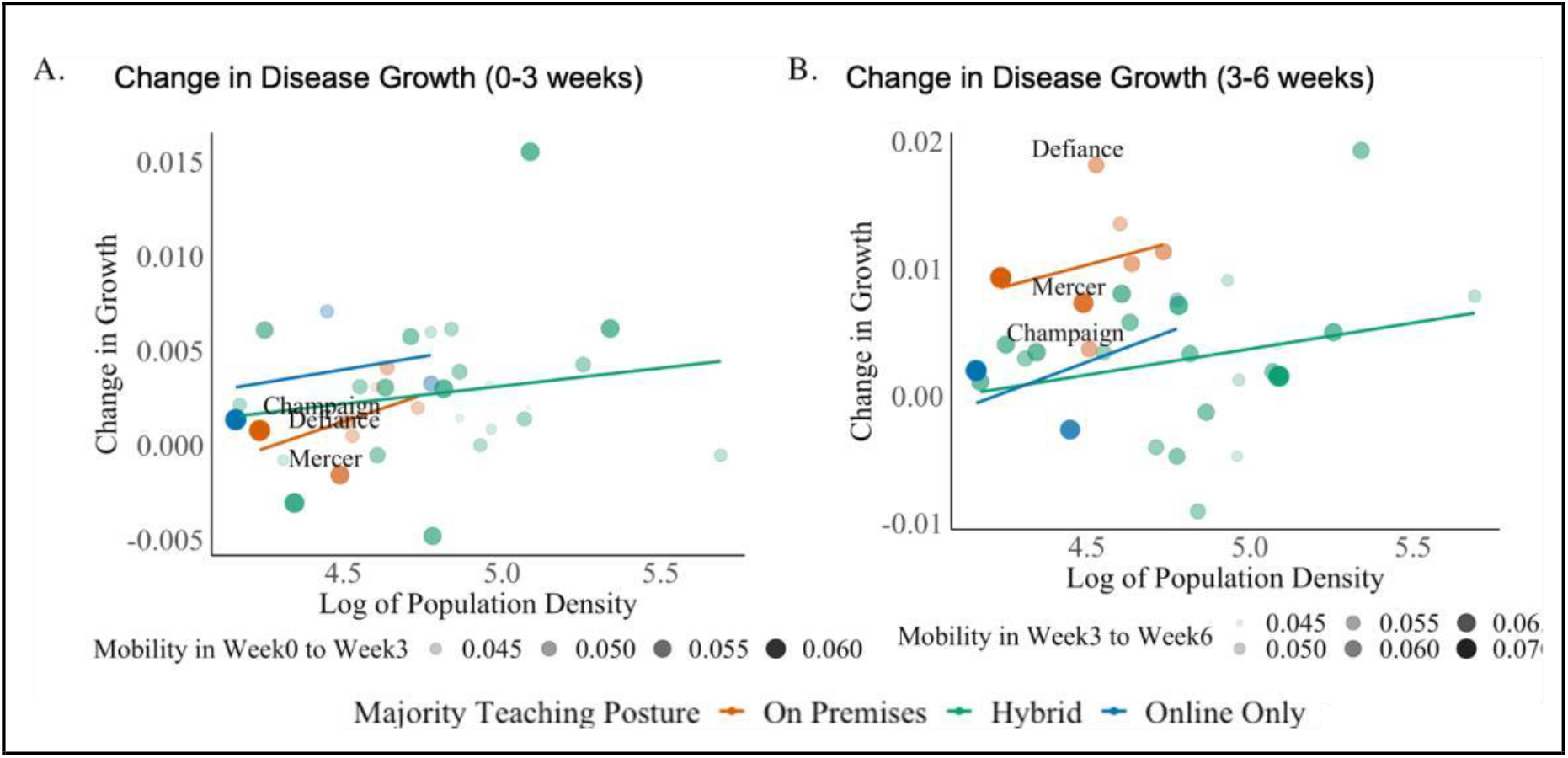
Change in growth of COVID-19 disease in micropolitan counties. For each micropolitan county the change in disease growth from A) the start of the fall semester to three weeks into the semester or B) from three weeks to six weeks into the fall semester. The average mobility of each county during these respective time periods is depicted by the changing circle diameters as indicated in the legend.

Fig 5B reveals some variation in COVID-19 disease growth during the fall semester for on-premises micropolitan counties, where some counties had much more severe change in growth compared to other counties (such as Defiance). In contrast counties such as Mercer and Champaign had COVID-19 disease growths similar to counties with online or hybrid learning (Fig 5B). Both Champaign and Defiance counties contain five school districts with similar percentages of students in hybrid (46-49%) and >50% in on-premises learning modes (Supplemental Table 1). In contrast, Mercer county had 89% of school age children in on-premises teaching but had one of the lowest COVID-19 growths during 3 to 6 weeks after the fall semester (Supplemental Table 1 and Fig 4B). Thus, the overall percentage of students within a county in on-premises learning does not directly correlate with an acceleration in disease growth. However, changes of NPI compliance and adherence to interventions could influence disease acceleration and was not captured in this study.

Ohio was chosen based on the implementation of universal state-wide interventions on masks orders, dining restrictions, and gathering bans. However, confounders such as adherence to interventions may be county specific and could not be captured within our data sets. In addition, it is unclear if students are functioning as the vector and spreading COVID-19 or whether teachers and school staff are the vehicles of SARS-CoV-2. Additionally, we cannot rule out that behaviors, other than mobility, in communities with predominantly on-premises teaching were different than those with predominantly online learning. These differences may also contribute to the spread of COVID-19. A recent study using self-reporting surveys conducted by the CMU Delphi group revealed that households with students in on-premises teaching had a higher odds ratio of COVID-19 like illness^9^. However, self-reported data may be inherently biased, and more information regarding infections in school age children is needed.

The analysis of Ohio counties based on teaching methods reveals a positive relationship between COVID-19 disease growth and on-premises teaching. Interestingly, counties with hybrid learning, while having similar cumulative deaths to on-premises counties at the end of the fall semester (Fig 1A), had better disease outcomes in micropolitan counties based on the change in disease growth during the fall semester (Fig 5B). Thus mitigation strategies such as decreased occupancy could impact the community burden of COVID-19. The increase of COVID-19 disease burden in on-premises counties was unlikely due to lack of general interventions as practices of mask usage among teachers and students was implemented by all school districts within Ohio in accordance with the Ohio Department of Health Director’s order effective Aug 14, 2020 (Supplemental material).

Importantly, Lesser et al^9^ used survey data to capture implementation of intervention strategies from masking, distancing, and extracurricular sports to suggest that some NPIs are more effective than others. In Ohio the vast majority of on-premises micropolitan Ohio school districts implemented similar interventions such as masking and distancing (Supplemental material). However, few details regarding air exchange and ventilation were available on the school district websites. In addition, it is unclear whether schools required health screenings to ensure that symptomatic children were kept home, this strategy could also help limit transmission.

Despite the positive correlation of on-premises education and COVID-19 disease growth in Ohio counties, we remain optimistic that schools can and should return with on-premises learning in the Fall of 2021. To mitigate transmission risk, we recommend implementation of interventions detailed in^10^. Briefly, we encourage the continued use of masks, increased air ventilation, air purifiers, as well as reduced occupancy within individual classrooms when feasible. In addition, vaccination of both teachers and pediatric populations will increase the barriers to transmission, which should increase the safety of reopening schools for on-premises learning. Ohio in particular has been creative with incentivizing vaccination among its residents with a vaccine lottery, where vaccines are registered for a monetary award, and at the time of writing this article ∼ 45% of all residents were fully vaccinated.

## Methods

### US school district COVID-19 interventions and status data

Data on the teaching status and COVID-19 interventions of each school district implemented in the Fall 2020 was obtained from MCH Strategic Data. COVID-19 impact: school district status—updates for fall 2020, accessed on Feb 2, 2021 (www.mchdata.com/covid19/schoolclosings). MCH data surveyed school districts around the country in Fall of 2020. Approximately 70% of school districts responded and include information of 35 different variables including the school district name, county, and teaching method (Full on premises, Hybrid/partial, remote learning only, unknown/undecided, Other, pending). Most counties in Ohio had information on the school districts within that county, the exception was Harrison and Vinton County, which were excluded from our analysis, because their school status information was missing. The data for school policies is at the individual school level. However, there can be multiple schools in a school district and multiple school districts in a county, which led us to aggregated the information about the schools to the county level. The variable of most interested in is “*Majority_teaching_posture*”; which was calculated based on the proportion of students in each teaching mode within a given county.

### COVID-19 death data

Death data per county per day was obtained from data compiled by the Johns Hopkins Center for Systems Science and Engineering (JHU-CSSE) COVID-19 resource repository available at: github.com/CSSEGISandData/COVID-19.

### Mobility data

Mobility data was obtained from covidcast program within the DELPHI group at Carnegie Melon University (https://delphi.cmu.edu/covidcast/) in the SafeGraph COVIDcast epidata API for assessing social distancing metrics. Data was updated until April 19, 2021. This data source uses anonymized location data from mobile phones to measure mobility. The data is provided for individual census block groups with differential privacy at aggregated county or state levels. The data set contains 9 variables regarding mobility within a county or state. In our analysis variable ‘*full_time_work_prop*’ was used to infer the mobility within Ohio counties, and provides the fraction of mobile devices that spent more than 6 hours at a location other than their home zipcode during the daytime. The earliest data available is January 1, 2019. Safegraphs’s measures mobility each day which will produce biases on the weekend, where there is considerable more mobility, than on the weekday.

### County Level Covariates

Data for county population was obtained in the R_package ‘USmap’ which contains 2015 census data. Other county level covariates such as percent of 65+ population and uninsured population was obtained by the CDC Data Tracker “Trends in COVID-19 Cases and Deaths in the United States, by County-level Population Factors” available (https://covid.cdc.gov/covid-data-tracker/#pop-factors_7daynewcases).

## Data Availability and Code Availability

Details on model describing COVID-19 disease is included in Supplemental material. Our analysis is based on four publicly available datasets listed below:

1. Ohio K12 data Data source: https://www.mchdata.com/covid19/schoolclosings
2. County-level daily deaths and cases from COVID-19 in Ohio State Source: John Hopkins COVID Resource (https://github.com/CSSEGISandData/COVID-19)
3. County-level population mobility (SafeGraph data from COVID-CAST API created by CMI DELPHI group). Data source: https://delphi.cmu.edu/covidcast/
4. County-level demographic information of Ohio State (Ohio profile data from CDC). Data source: https://data.cdc.gov and https://covid.cdc.gov/covid-data-tracker/#pop-factors_7daynewcases

Cleaned and combined data sets, as well as the code for our generative model is available at https://github.com/alexazhu/36726-PIGHT-COVID.git

## Supporting information

Technical Appendix of Mathematical Model

## Data Availability

All data is publicly available at these sources:
1. Ohio K12 data Data source: https://www.mchdata.com/covid19/schoolclosings
2. County-level daily deaths and cases from COVID-19 in Ohio State Source: John Hopkins COVID Resource (https://github.com/CSSEGISandData/COVID-19)
3. County-level population mobility (SafeGraph data from COVID-CAST API created by CMI DELPHI group). Data source: https://delphi.cmu.edu/covidcast/
4. County-level demographic information of Ohio State (Ohio profile data from CDC). Data source: https://data.cdc.gov and https://covid.cdc.gov/covid-data-tracker/#pop-factors_7daynewcases
All our code and repositories are available in the data availability link below.

https://github.com/alexazhu/36726-PIGHT-COVID.git

## Acknowledgements

We thank Dr. Carter Mecher of the Public Health Company for useful discussions and members of the Lakdawala lab for critical review of this manuscript. SSL is funded by National Institute of Allergy and Infectious Diseases (CEIRS HHSN272201400007C).

## Author contributions

CE, YL, ZY, ZZ, VV performed the data analysis, SD and AA collected data, JL, RN, VV, SL conceived of the project, RN, VV, SSL wrote and edited the manuscript.

## Supplemental Figures

**Supplemental Table 1:** Breakdown of teaching mode percentiles within Micropolitan Counties

| Metropolitan County | %of students within each teaching posture |  |  | Primary Teaching |
| --- | --- | --- | --- | --- |
|  | Hybrid | On-premises | Online |  |
| ASHLAND | 73% | 0% | 13% | Hybrid |
| ASHTABULA | 63% | 26% | 11% | Hybrid |
| ATHENS | 64% | 0% | 36% | Hybrid |
| AUGLAIZE | 21% | 79% | 0% | On_Premises |
| CHAMPAIGN | 46% | 54% | 0% | On_Premises |
| CLINTON | 40% | 23% | 37% | Hybrid |
| COLUMBIANA | 51% | 12% | 20% | Hybrid |
| COSHOCTON | 100% | 0% | 0% | Hybrid |
| CRAWFORD | 0% | 54% | 46% | On_Premises |
| DARKE | 33% | 24% | 43% | Online_Only |
| DEFIANCE | 49% | 51% | 0% | On_Premises |
| ERIE | 76% | 11% | 0% | Hybrid |
| FAYETTE | 100% | 0% | 0% | Hybrid |
| GALLIA | 0% | 0% | 47% | Online_Only |
| GUERNSEY | 100% | 0% | 0% | Hybrid |
| HANCOCK | 95% | 5% | 0% | Hybrid |
| HURON | 61% | 27% | 0% | Hybrid |
| JACKSON | 47% | 0% | 30% | Hybrid |
| KNOX | 0% | 23% | 32% | Online_Only |
| LOGAN | 37% | 63% | 0% | On_Premises |
| MARION | 52% | 0% | 0% | Hybrid |
| MERCER | 0% | 89% | 11% | On_Premises |
| MUSKINGUM | 100% | 0% | 0% | Hybrid |
| OTTAWA | 65% | 35% | 0% | Hybrid |
| ROSS | 48% | 41% | 11% | Hybrid |
| SANDUSKY | 44% | 0% | 0% | Hybrid |
| SCIOTO | 74% | 0% | 26% | Hybrid |
| SENECA | 69% | 6% | 0% | Hybrid |
| SHELBY | 77% | 23% | 0% | Hybrid |
| TUSCARAWAS | 80% | 6% | 14% | Hybrid |
| VAN WERT | 18% | 82% | 0% | On_Premises |
| WASHINGTON | 88% | 12% | 0% | Hybrid |
| WAYNE | 56% | 25% | 0% | Hybrid |

**Supplemental Figure 1.**
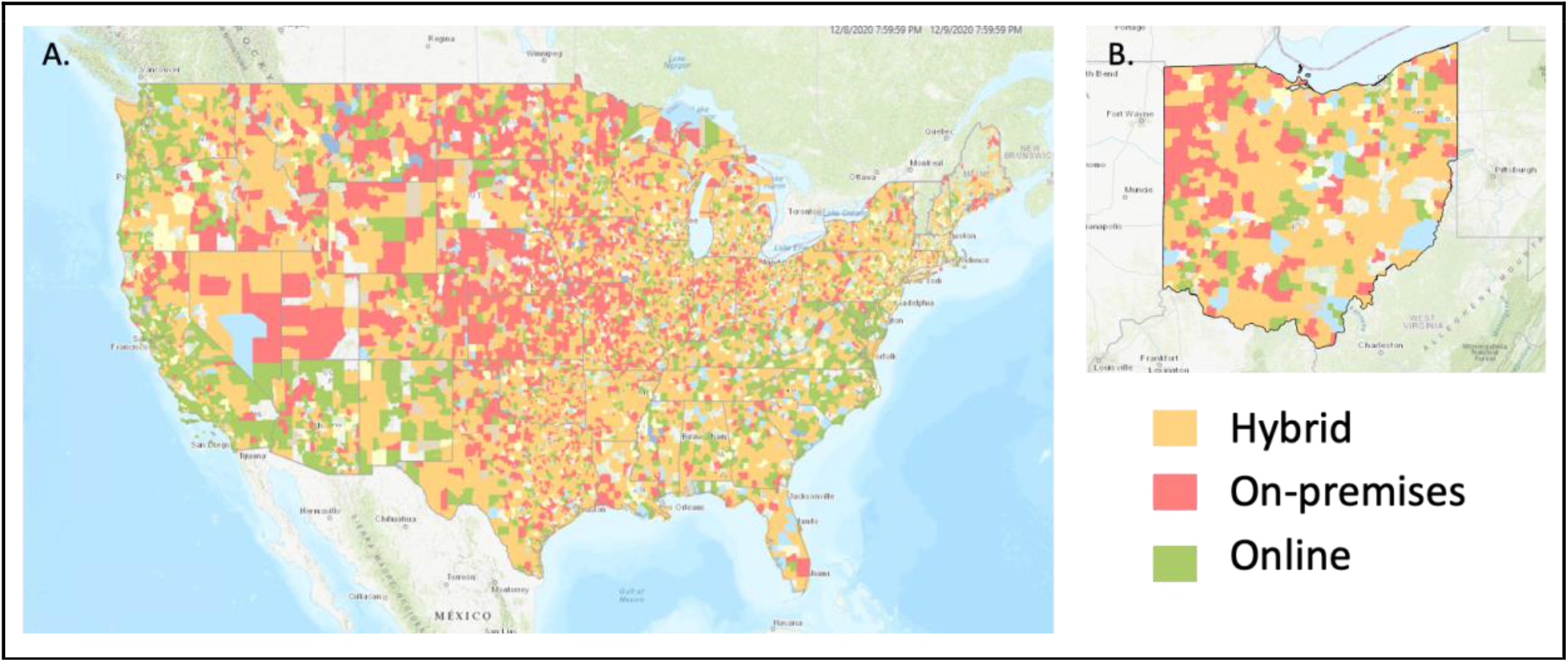
Teaching posture for each school district across the United States and within a state varied between on-premises, online only, and hybrid. Survey data of all school districts across the United States provided by MCH data (https://www.mchdata.com/covid19/schoolclosings). A) depicts the continuous United States. B) A map of Ohio School Districts with available teaching posture by school district reveals the variation across the state.

**Supplemental Figure 2.**
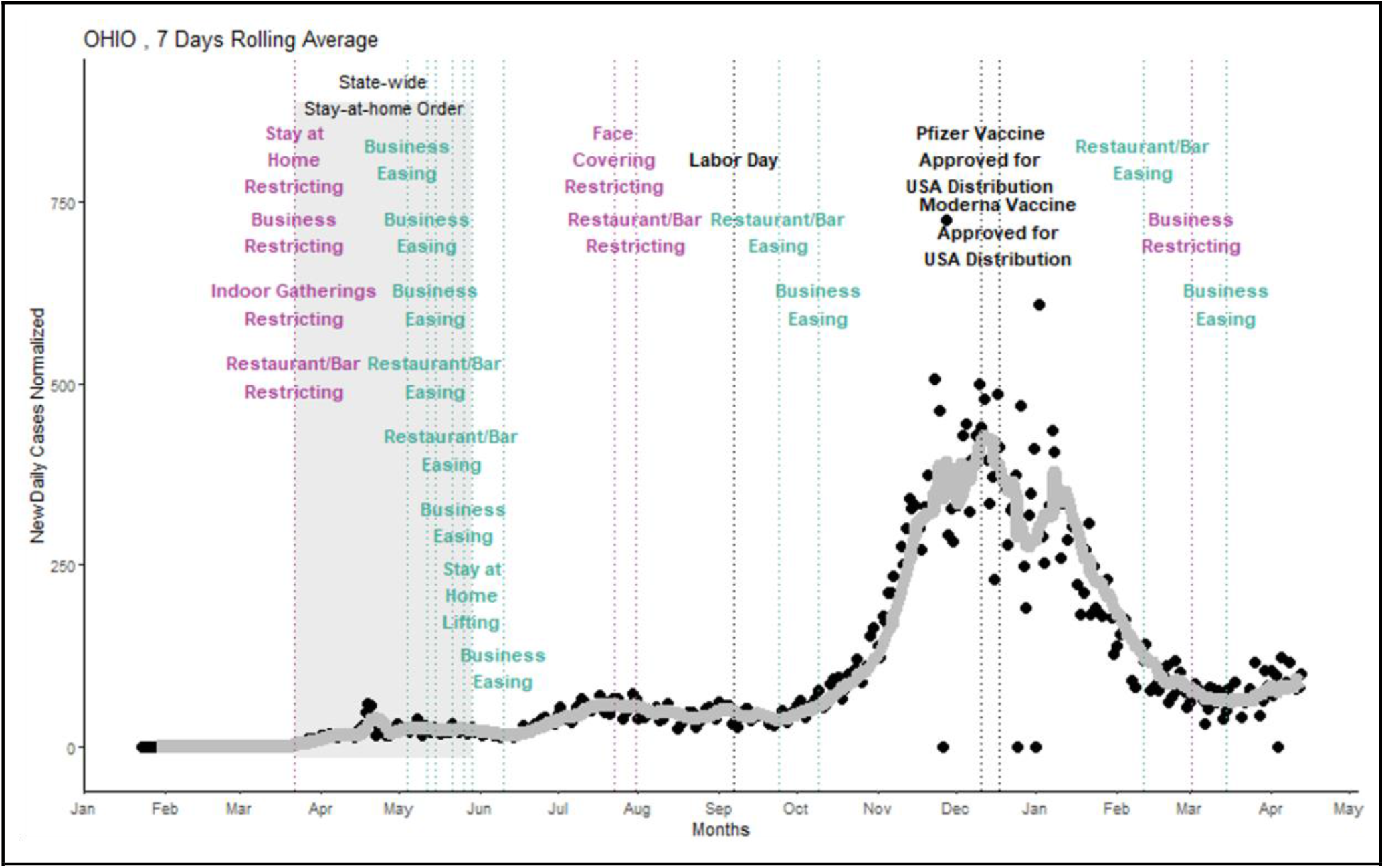
Trajectory of COVID-19 cases in Ohio and implementation of statewide interventions. Daily COVID-19 case increases are presented in black circles and a seven day rolling average (gray line) portray the trajectory of the pandemic in Ohio. State level interventions where implemented throughout the 2020 and 2021. These include implementation of restrictions on restaurant and bars, gathering size limits, business restrictions, and mask order. Each intervention is listed and is color coded for magenta – restriction and teal – easing. The initial stay at home order place in March of 2020 was lifted in June of 2020 and is indicated by the gray shaded box. An interactive version of this graph can be found at www.PHIGHTCOVID.org

**Supplemental Figure 3.**
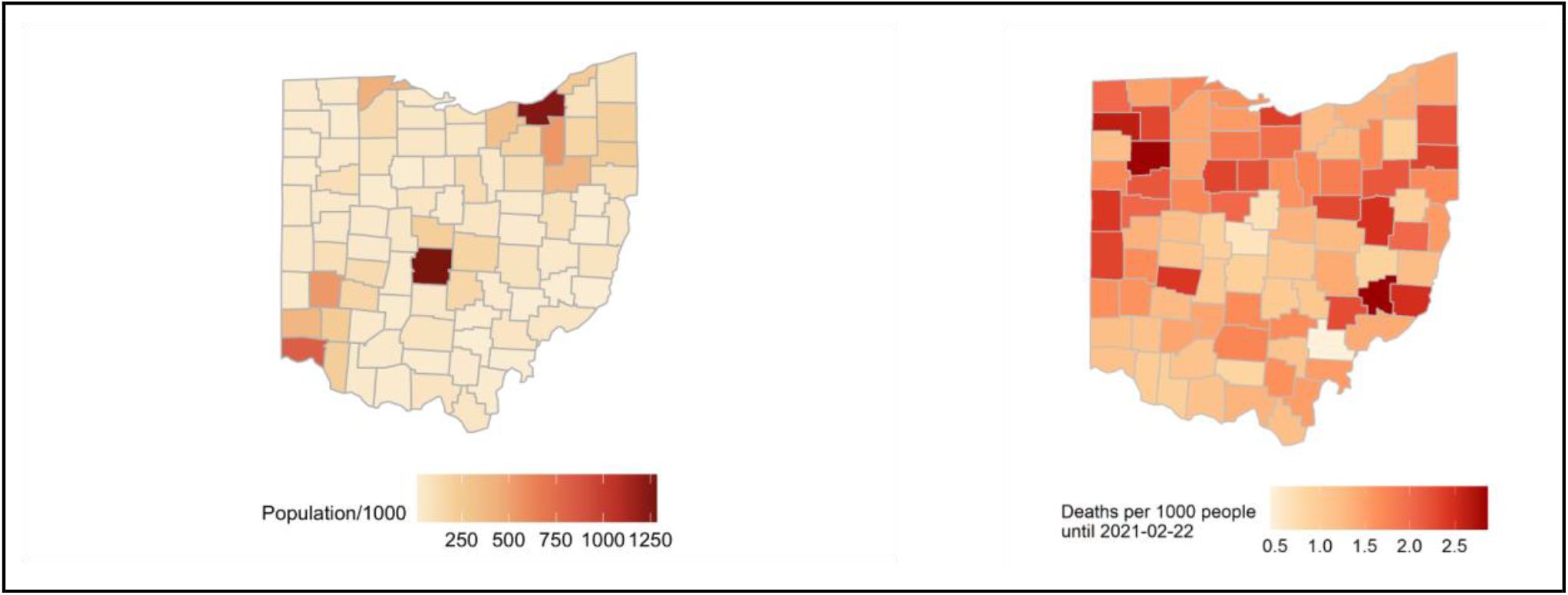
Population density and COVID-19 deaths in Ohio by county. A) Population density (data from 2015 Census in R package ‘USmap’) of each county per 1000 individuals. B) cumulative COVID-19 deaths (data from JHU Coronavirus resource center) from March 2020 - Feb 22, 2021 per 1000 people.

**Supplemental Figure 4.**
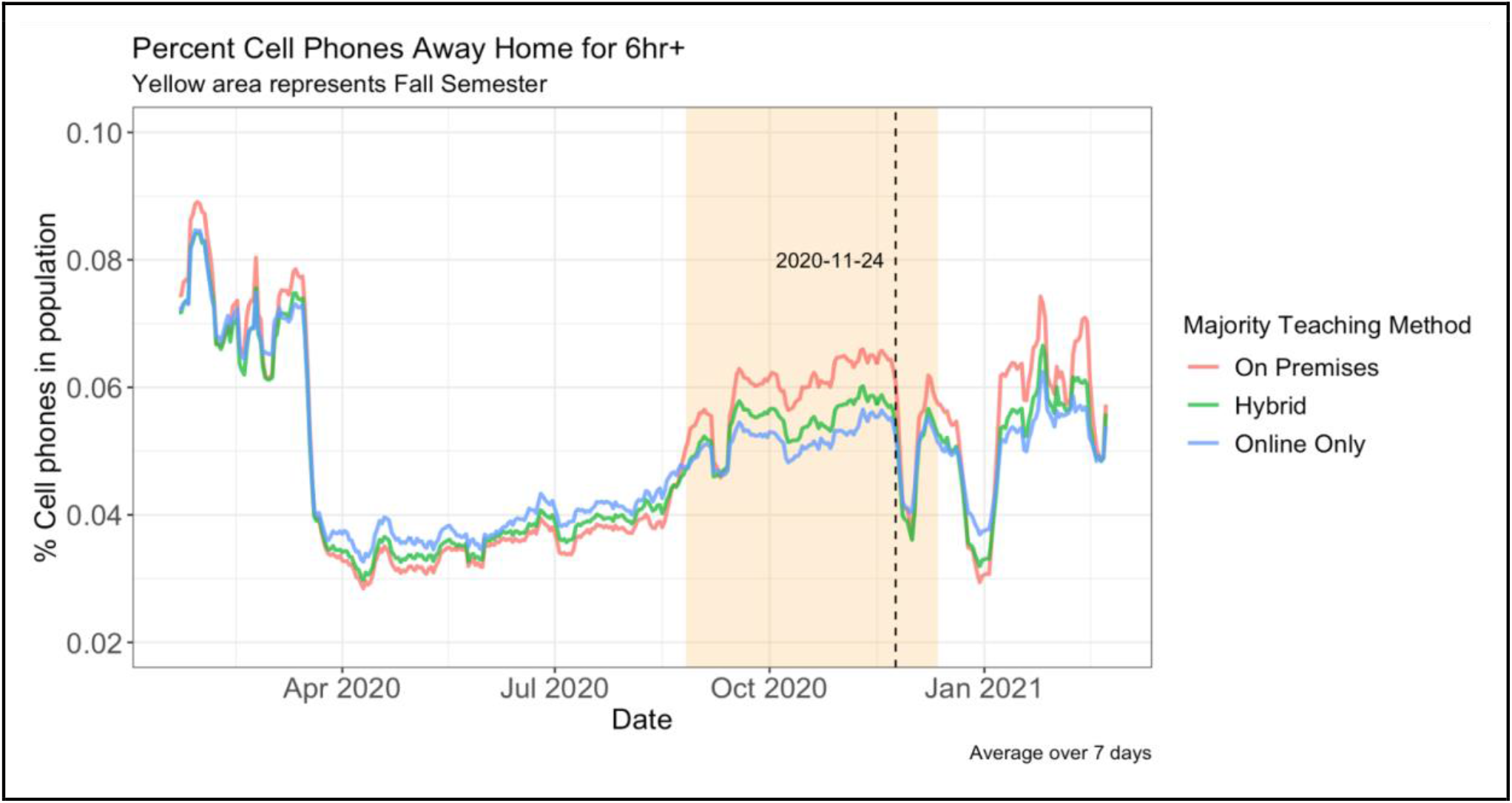
Mobility of cell phones during the COVID pandemic in counties based on teaching posture. COVIDcast collates data on mobility based on movement of cellphones out of the home zipcode for longer than 6 hours. The average mobility is presented for all counties in a specified teaching group from March 2020 to April 2021. The yellow shaded area indicates the fall semester.

**Supplemental Figure 5.**
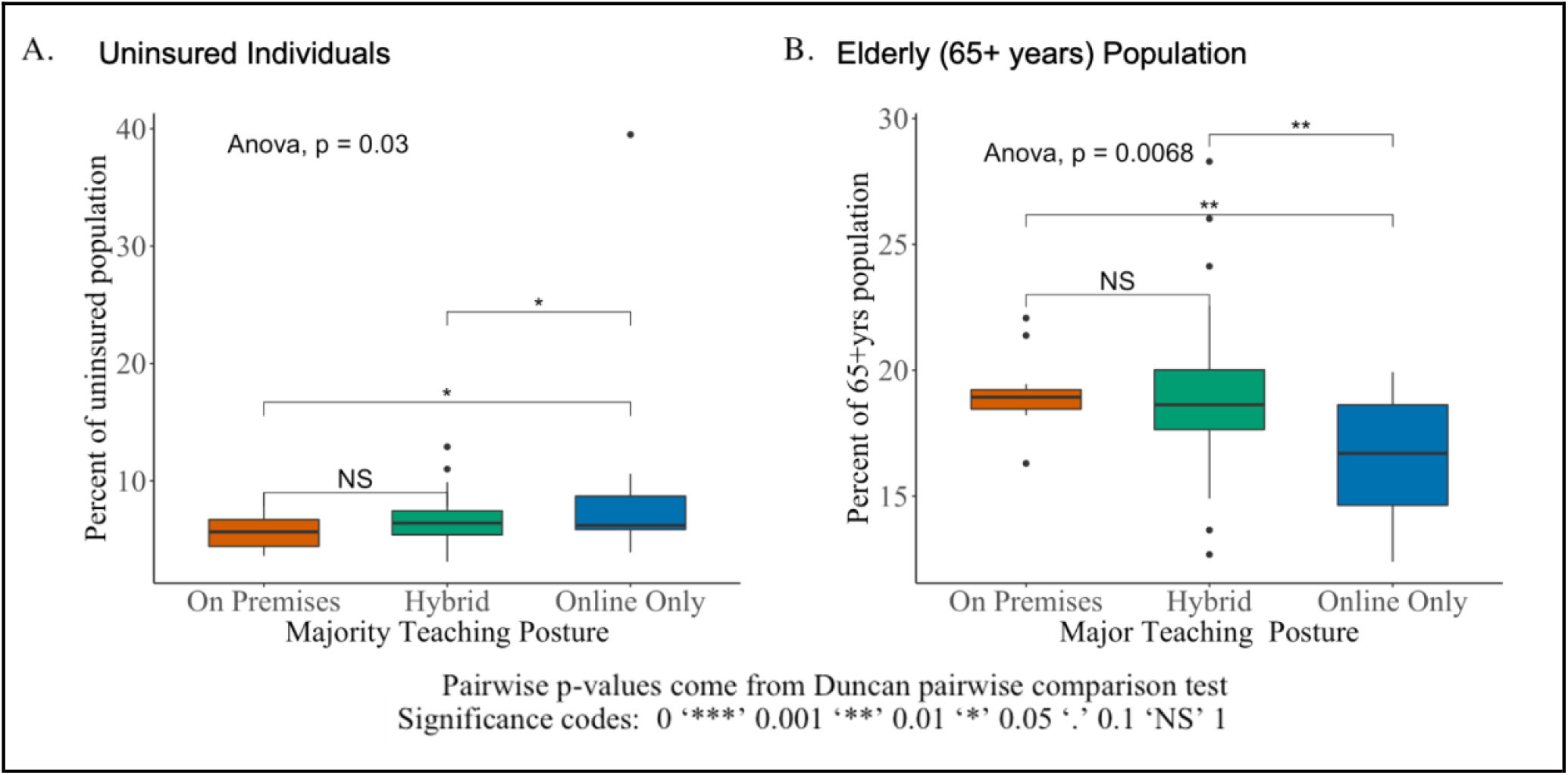
Breakdown of uninsured individuals and elderly in counties based on teaching posture. A) Percent of uninsured individuals in each county and B) percent of individuals over 65 years of age per county were obtained for Ohio micropolitan counties from CDC ‘COVID Data tracker by county-level population factors’ (https://covid.cdc.gov/covid-data-tracker/#pop-factors_7daynewcases). Statistical comparisons were performed using ANOVA and Duncan pairwise comparison.

**Supplemental Figure 6.**
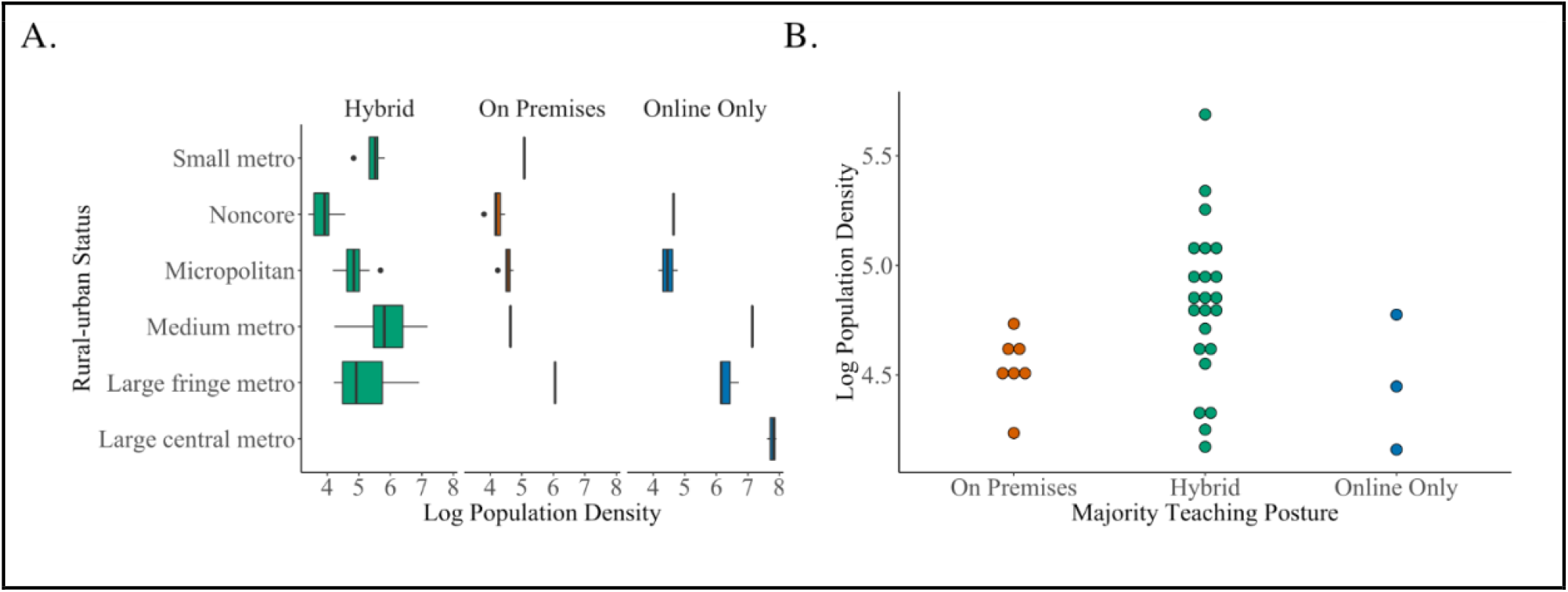
Breakdown of Urban/Rural counties based on the primary teaching posture. A) Each Ohio county is broken down by its specific urban/rural status from large central metro to small metro and as log population density. B) A scatter plot of the micropolitan counties based on log population density, each spot is an individual county.

## References

1 Falk, A. et al. COVID-19 Cases and Transmission in 17 K-12 Schools - Wood County, Wisconsin, August 31-November 29, 2020. MMWR Morb Mortal Wkly Rep 70, 136–140, doi:10.15585/mmwr.mm7004e3 (2021).

2 Ismail, S. A., Saliba, V., Lopez Bernal, J., Ramsay, M. E. & Ladhani, S. N. SARS-CoV-2 infection and transmission in educational settings: a prospective, cross-sectional analysis of infection clusters and outbreaks in England. Lancet Infect Dis 21, 344–353, doi:10.1016/S1473-3099(20)30882-3 (2021).

3 Cao, Y., Hiyoshi, A. & Montgomery, S. COVID-19 case-fatality rate and demographic and socioeconomic influencers: worldwide spatial regression analysis based on country-level data. BMJ Open 10, e043560, doi:10.1136/bmjopen-2020-043560 (2020).

4 Glezen, W. P. & Couch, R. B. Interpandemic influenza in the Houston area, 1974-76. N Engl J Med 298, 587–592, doi:10.1056/NEJM197803162981103 (1978).

5 Team, C. C.-R. Geographic Differences in COVID-19 Cases, Deaths, and Incidence - United States, February 12-April 7, 2020. MMWR Morb Mortal Wkly Rep 69, 465–471, doi:10.15585/mmwr.mm6915e4 (2020).

6 Verity, R. et al. Estimates of the severity of coronavirus disease 2019: a model-based analysis. Lancet Infect Dis 20, 669–677, doi:10.1016/S1473-3099(20)30243-7 (2020).

7 Statistics, N. C. f. H. NCHS Urban-Rural Classification Scheme for Counties, <https://www.cdc.gov/nchs/data_access/urban_rural.htm> (2013).

8 Hernan, M. A., Clayton, D. & Keiding, N. The Simpson’s paradox unraveled. Int J Epidemiol 40, 780–785, doi:10.1093/ije/dyr041 (2011).

9 Lessler, J. et al. Household COVID-19 risk and in-person schooling. Science 372, 1092–1097, doi:10.1126/science.abh2939 (2021).

10 Richard Corsi, S. L. M., Marissa G. VanRy, Linsey C. Marr, Leslie R. Cadet, Nira R. Pollock, David Michaels, Emily R. Jones, Meira Levinson, Yuguo Li, Lidia Morawska, John Macomber, Joseph G. Allen. Designing infectious disease resilience into school buildings through improvements to ventilation and air cleaning. THE LANCET COVID-19 COMMISSION (2021).

