## Supplementary material for "School Teaching Posture Correlates with COVID-19 Disease Outcomes in Ohio": Technical Appendix of Mathematical Model

### The model

Below we describe a two stage observation model for the time series of deaths. The aim is to fit this model the observed deaths to infer $B_t$, the growth factor of the disease at time $t$, for all $t$.

Let $I_t$ denote *new* infections in day $t$ and $Y_t$ deaths in day $t$. Note that $Y_t$ is observed but $I_t$ is not observed; in statistical jargon, $I_t$ is a latent variable. Given new infections $I_{t-1}$ at time $t-1$, let

$$I_t = I_{t-1}e^{B_t} + \delta_t \tag{1}$$

where $\delta_t$ are mean 0 random variables (independent of all other variables) and $B_t$ measures the transmission of the epidemic at time $t$. This parameter is particularly interpretable: if $B_t < 0$, then $e^{B_t} \in [0, 1)$, meaning that the number of new daily infections is decreasing at time $t$: the disease is shrinking. Conversely, if $B_t \geq 0$, then $e^{B_t} \geq 1$, meaning that the disease is growing. The growth of the epidemic varies across time as the number of susceptible decreases over time (assuming re-infections are unlikely), various interventions take effect, the weather changes, etc.

Next, given new infections up to $t$, we express deaths at $t$ as

$$Y_t = \sum_{s=1}^{t} f(s,t)I_s + \xi_t \tag{2}$$

where $\xi_t$ are mean 0 random variables (independent of the other variables) and $f(s,t)$ denotes the probability that someone infected at time $s$ dies at time $t$. We further write

$$f(s,t) = d(s)f_0(s,t) \tag{3}$$

where $d(s)$ is the probability that someone infected at time $s$ will eventually die and $f(s,t)$ is the probability that someone infected at time $s$ and who will eventually die, will die at time $t$. Following Bonvini et al. (2021), we take $f_0(s,t)$ to be a Gamma distribution with mean 23.9 days and coefficient of variation 0.40.

Finally, we recurse (1) and plug in (2) to obtain the mean model for deaths

$$\mathbb{E}[Y_t] = \sum_{s=1}^{t} I_1 f(s,t) e^{\sum_{r=1}^{s} B_r}. \tag{4}$$

**Simplified Model** The non-linear form of (4) makes it difficult to estimate $B_t$, the growth of the disease. We therefore simplify $f_0(s,t)$ in (3) to be a point mass at its mean, $\delta = 24$ days, and obtain the simplified model:

$$\mathbb{E}[Y_t] = I_1 d(t-\delta) e^{\sum_{r=1}^{t-\delta} B_r}.$$

Now, taking the log, we get $\mathbb{E}[\log(Y_t + 1)]$[1] $\approx \log[d(t-\delta)] + \log I_1 + \sum_{r=1}^{t-\delta} B_r$, or equivalently

$$\mathbb{E}[\log(Y_{t+\delta} + 1)] \approx \log[d(t)] + \log I_1 + \sum_{r=1+\delta}^{t} B_r. \tag{5}$$

This model is additive and thus easily fit to data, as shown in the following plot for the times series of deaths in the three majority teaching methods groups of counties. The curves are highly non-linear so we fitted a local linear smoother with bandwidth 4 weeks.

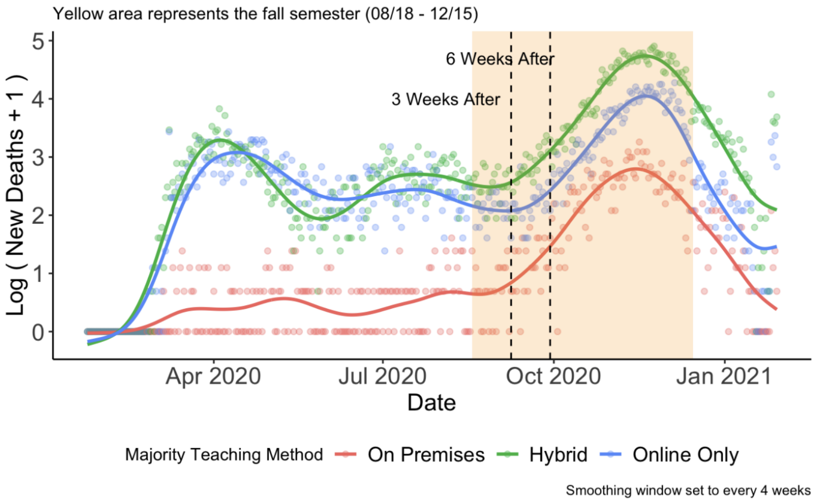


The last step is to notice that the derivative of (5) is

$$\partial\mathbb{E}[\log(Y_{t+\delta} + 1)]/\partial t = d'(t)/d(t) + B_t,$$

meaning that we can estimate $d'(t)/d(t) + B_t$ by taking the derivatives with respect to time of the curves shown above. These derivatives are plotted in Fig. 2B. The probability of dying, $d(t)$, certainly changed over time, e.g. hospitals were better prepared during the second wave. They may also vary across counties, e.g. be higher in older counties. However, it is reasonable to assume that, within each county, $d(t)$ is constant during the fall semester, in which case

$$\partial\mathbb{E}[\log(Y_{t+\delta} + 1)]/\partial t = B_t.$$

We are able to estimate the growth of the disease even though we did not observe the number of infections.

[1] We add 1 in case $Y_t = 0$.

### References

Bonvini, M., Kennedy, E., Ventura, V. and Wasserman, L. (2021). Causal Inference in the Time of Covid-19.
