## Supplemental material for "School Teaching Posture Correlates with COVID-19 Disease Outcomes in Ohio"

Ohio | Department of Health

Mike DeWine, Governor
Jon Husted, Lt.Governor

Lance D. Himes, Interim Director

### DIRECTOR'S ORDER

#### Re: Director's Order Requiring the Use of Facial Coverings in Child Education Settings

I, Lance D. Himes, Interim Director of the Ohio Department of Health (ODH), pursuant to the authority granted to me in R.C. 3701.13 to "make special orders…for preventing the spread of contagious or infectious diseases" **Order** the following to prevent the spread of COVID-19 into the State of Ohio:

1. **Facial Coverings (Masks).** Except as provided herein, all students, faculty, and staff in any child care setting, school building, or other location that provides care or education to any child in kindergarten through grade twelve in the State of Ohio shall wear facial coverings at all times when:

    a. In any indoor location including, but not limited to, classrooms, gymnasiums, offices, locker rooms, hallways, cafeteria, and/or locker bays;

    b. Outdoors on school property and unable to consistently maintain a distance of six feet or more from individuals who are not members of their household;

    c. Waiting for a school bus outdoors and unable to maintain a distance of six feet or more from individuals who are not members of their household; or

    d. Riding a school bus.

    For purposes of this Order, a facial covering (mask) is any material that covers an individual's nose, mouth and chin.

2. **Exemptions.** The requirement to wear a facial covering does not apply when:

    a. The individual has a medical condition including respiratory conditions that restricts breathing, mental health conditions, or a disability that contraindicates the wearing of a facial covering; or

    b. The individual is communicating or seeking to communicate with someone who is hearing impaired or has another disability, where an accommodation is appropriate or necessary;

    c. The individual is actively participating in outdoor recess and/or physical activity where students are able to maintain a distance of six feet or more or athletic practice, scrimmage, or competition that is permitted under a separate Department of Health Order;

    d. The individual is seated and actively consuming food or beverage;

    e. Where students and staff can maintain distancing of at least six feet and removal of the facial covering is necessary for instructional purposes, including instruction in foreign language, English language for non-native speakers, and other subjects where wearing a facial covering would prohibit participation in normal classroom activities, such as playing an instrument;

f. Students are able to maintain a distance of six feet or more and a mask break is deemed necessary by the educator supervising the educational setting;

g. The individual is alone in an enclosed space, such as an office; or

h. When an established sincerely held religious requirement exists that does not permit a facial covering.

3. **Duration:** This Order shall be effective at 12:01 a.m. on August 14, 2020 and remains in full force and effect until the State of Emergency declared by the Governor no longer exists or the Director of the Ohio Department of Health rescinds or modifies this Order, whichever occurs sooner.

4. **Conflicting Orders:** The requirements of this Order are controlling and effective over any conflicting provisions of the "Director's Order for Facial Coverings throughout the State of Ohio", issued on July 23, 2020.

Accordingly, I hereby **ORDER** that all persons wear facial coverings subject to the circumstances and requirements of this Order. This Director's Order shall remain in full force and effect until the State of Emergency declared by the Governor no longer exists, or the Director of the Ohio Department of Health rescinds or modifies this Order. To the extent any public official enforcing this Director's Order has questions regarding the effect of this Order, the Director of Health hereby delegates to local health departments the authority to answer questions in writing and consistent with this Order.

Lance D. Himes
Interim Director of Health

August 13, 2020

### Ohio Micropolitan Counties School Districts COVID-19 Reopening Plans

### School District Sources

| County | School District | Sources |
|---|---|---|
| Auglaize | Auglaize County Educational Academy | https://www.aceva.org/WhyACE.aspx |
| Auglaize | Minster Local | https://docs.google.com/document/d/1AcyJaJGcafcMxHKAga3n5NICSOHTwoildvSKP21zx9Y/edit |
| | | https://www.minsterschools.org/about-us/athletics/ |
| Auglaize | New Bremen Local | https://www.newbremenschools.org/Downloads/2020NBReopeningPlan.pdf |
| | | https://twitter.com/newbremenschool?lang=en |
| | | https://www.bremen.k12.oh.us/protected/mastercalendar.aspx |
| Auglaize | New Knoxville Local | http://www.nk.k12.oh.us/wp-content/uploads/2020/08/NK-Face-Covering-Plan.pdf |
| | | http://www.nk.k12.oh.us/wp-content/uploads/2020/07/20200723075843665.pdf |
| | | http://www.nk.k12.oh.us/wp-content/uploads/2020/08/2020-2021-Calendar.pdf |
| Auglaize | St. Mary's City | https://www.smriders.net/RiderRestart.aspx |
| | | https://www.facebook.com/page/192207320966733/search/?q=basketball |
| Auglaize | Wapakoneta City | https://www.wapak.org/Downloads/Wapakoneta%20Guidelines%207_8_20.pdf |
| | | https://www.facebook.com/WapakCitySchool/posts/4776159695757669 |
| | | https://www.facebook.com/WapakCitySchool/posts/4397710323602610 |
| | | https://www.facebook.com/page/511278345579180/search/?q=football |
| Auglaize | Waynesfield-Goshen Local | https://docs.google.com/document/d/1jrsCGeX2x6w6etve5IkBXnaSD2N-Bc-L1ZvgBZYws1c/edit |
| | | https://core-docs.s3.amazonaws.com/documents/asset/uploaded_file/800258/Calendar_Schedule_2020-21__Public_.pdf |
| | | https://www.wgschools.org/o/wga |
| Champaign | Graham Local | https://drive.google.com/drive/folders/1KTMPiAmwd1bJJI9WEHh3LGpsywjH4rTC |
| | | https://filecabinet5.eschoolview.com/CA7BA98B-DAE2-42EE-BC3A-AA7201E54C54/466b610d-a297-41f0-8753-5e56b8fcdff6.pdf |
| | | https://grahamathletics.org/teams/2929273/boys/football/varsity |
| Champaign | Mechanicsburg Exempted Village | https://docs.google.com/document/d/12spdR1NMd0fnmSrUaT8Q5zOsCk8LdBVoUNd2f8_CCKw/edit?fbclid=IwAR1sb-19bLML8eyBLyxDBeX1DoDisP-PGRg8PT6kVLsYe9olrXLsS3VBbbQ |
| | | https://www.facebook.com/McBurg.Schools/posts/730059610941993 |
| | | https://www.mcburg.org/o/athletics |
| | | https://docs.google.com/document/d/1QboZZ4LV21HafPP8gyfsNaK8YE9kacTdNMlMSX78Jps/edit?fbclid=IwAR046gQnE7oyU0mlLExfcl9Vp5lpfA9Ylm4IQBf9__uTCn7aQWjj6xW7lY |
| Champaign | Triad Local | https://drive.google.com/drive/folders/1zeSxAjWQ7lW29F4jIu8gOfeT-tyzYF5t |
| | | https://www.triadk12.org/calmonthview.aspx?schoolid=0&t=&schoollistids=#m=3&s=0&t=1:2:24&y=2021 |
| | | https://www.facebook.com/page/727438614045834/search/?q=hybrid |
| Champaign | Urbana City | https://www.urbanacityschools.org/docs/district/forms&documents/hillclimber%20reset%20and%20restart%20plan%20public%20communication%207-22-20%20-%20google%20docs.pdf?id=1290 |
| | | https://www.urbanacityschools.org/docs/building/1/newsletters/2020-21/hsnewsletter120420.pdf?id=1482 |
| Champaign | West Liberty-Salem Local | https://core-docs.s3.amazonaws.com/documents/asset/uploaded_file/814948/FAQs_2020-21_WL-S_School_Year.pdf |
| | | https://core-docs.s3.amazonaws.com/documents/asset/uploaded_file/918804/2020-2021_WLS_District_Calendar_V4.pdf |

| | | |
|---|---|---|
| | | https://www.facebook.com/permalink.php?story_fbid=858392185004419&id=375356183308024 |
| Crawford | Buckeye Central Local | https://www.buckeye-central.k12.oh.us/3/News/1236#sthash.DeHYfv1r.dpbs |
| | | https://www.facebook.com/page/697777360302561/search/?q=remote |
| | | https://www.facebook.com/page/697777360302561/search/?q=basketball |
| | | https://www.buckeye-central.org/userfiles/4/my%20files/late%20start%20days%202020-2021%201st%20semester.pdf?id=4532 |
| Crawford | Bucyrus City | https://www.bucyrusschools.org/userfiles/2/my%20files/crawford%20county%202020%20copy.pdf?id=776 |
| | | https://www.bucyrusschools.org/2/content/athletics |
| | | https://www.bucyrusschools.org/userfiles/2/my%20files/2020-2021%20cal%20revised.pdf?id=700 |
| | | https://www.facebook.com/page/146425926688/search/?q=remote |
| Crawford | Colonel Crawford Local | https://www.cck12.org/images/2020-CC-COVID-19-Reopening-Guidance.pdf |
| | | https://www.facebook.com/page/101794504953281/search/?q=remote |
| | | https://www.facebook.com/colonelcrawfordlocalschools/posts/131385891994142 |
| Crawford | Crestline Exempted Village | https://www.crestlinebulldogs.org/blog/2020/11/12/in-person-classes-resume |
| | | https://www.crestlinebulldogs.org/upload/remote_learning_update_12_6_2020.pdf |
| | | https://www.crestlinebulldogs.org/athletics-2 |
| | | https://www.crestlinebulldogs.org/upload/final_crestline_restart_fall_2020_2.pdf |
| | | https://www.crestlinebulldogs.org/blog/2020/10/27/remote-learning-update-10272020 |
| Crawford | Galion City | https://www.galionschools.org/upload/documents/20-21_school_year/galion-013_reopening_plan_2.pdf |
| | | https://www.facebook.com/galioncityschools/posts/1423282364541187 |
| | | https://www.facebook.com/page/303963359806432/search/?q=football |
| | | https://www.facebook.com/galioncityschools/posts/1411300009072756 |
| Crawford | Wynford Local | https://arbiterlive.com/School/Calendar/26327 |
| | | https://core-docs.s3.amazonaws.com/documents/asset/uploaded_file/826292/2020-07-30_-_Reopening_Plan.pdf |
| | | https://www.wynfordroyals.org/o/wynford/article/362694 |
| | | https://www.wynfordroyals.org/article/292320 |
| Defiance | Ayersville Local | http://ayersville.org/UserFiles/Servers/Server_43710/File/District_Forms/Restart%20Plan.pdf |
| | | https://www.facebook.com/page/159237007451993/search/?q=remote |
| | | http://www.ayersville.org/cms/one.aspx?pageId=43814 |
| Defiance | Central Local | https://www.centrallocal.org/apaches/files/Newsletters/DistrictNewsletter2020-08.pdf |
| | | https://www.centrallocal.org/apaches/files/News/FesParentInformationSheet2020-2021.pdf |
| | | https://www.facebook.com/fvapaches/posts/747758555977335 |
| | | http://www.ayersville.org/common/pages/UserFile.aspx?fileId=43628829 |
| | | https://www.facebook.com/page/138643953555468/search/?q=basketball |

| | | |
|---|---|---|
| Defiance | Defiance City | https://filecabinet.eschoolview.com/2E72D051-13D0-4789-91E9-A7D457D11578/c4fb5377-5c28-4bb0-ba21-f2471a189e23.pdf |
| | | https://www.facebook.com/page/432492390139521/search/?q=basketball |
| | | https://www.facebook.com/DefianceCitySchools/posts/3490203737701689 |
| Defiance | Hicksville Exempted Village | https://drive.google.com/file/d/1wt5hq3VVt-_iZWcST192iGmE65Cetr7i/view?fbclid=IwAR1A3lY9nYlkcOhG2FEofUTkoURNmAt854-lfBkvY8Yv4puax1MquOFqFS4 |
| | | https://drive.google.com/file/d/1D0b_GKzMn_gCFZAlfTaiV7lIMzOBgcB7/view?fbclid=IwAR2BDmqKlviL_0pNd_XGse4YfoA3rxJm3iSMubW4vHZrcDKvVBFA19LwpOI |
| | | https://drive.google.com/file/d/1Vx7ifMsFXiYv4J4XOAv69CKGEiLprt9s/view?fbclid=IwAR2Xd6aZVOsiZvAVisuZBF_iS1oEnfDDvO5iXm2ie9ZSYCccRGGfSr7sg88 |
| | | https://www.facebook.com/page/171365296239720/search/?q=football |
| Defiance | Northeastern Local | https://core-docs.s3.amazonaws.com/documents/asset/uploaded_file/815084/REOPENING_TINORA_2020__9_.pdf |
| | | https://core-docs.s3.amazonaws.com/documents/asset/uploaded_file/1045913/High_School_Remote_Learning_2020-2021.pdf |
| | | https://www.tinora.org/athletics?filter_ids=45544 |
| Logan | Bellefontaine City | http://www.bellefontaine.k12.oh.us/district/public_annoucements/reopening_information |
| | | http://www.bellefontaine.k12.oh.us/district/public_annoucements/back_to_school_information |
| | | http://www.bellefontaine.k12.oh.us/district/public_annoucements/back_to_school_parent_info |
| | | https://bellefontaineathletics.com/events |
| Logan | Benjamin Logan Local | https://benjaminloganathletics.com/main/calendar/ |
| Logan | Indian Lake Local | http://www.ils-k12.org/laker_news/indian_lake_schools_reopening_plan___faqs |
| Logan | Riverside Local | https://www.painesville-township.k12.oh.us/remotelearningacademy_home.aspx |
| | | https://www.painesville-township.k12.oh.us/SportSchedules.aspx |
| Mercer | Coldwater Exempted Village | https://drive.google.com/file/d/1DVU25BVcl82bX2CB0v_9K45N2P-J78Oz/view?fbclid=IwAR1fl6KuBO9mhfe-wvw6fkMNb_X35kwO3K4RExr-BDD1vTFZr2p4lZFpElk |
| Mercer | Parkway Local | http://www.parkwayschools.org/community_news |
| | | https://docs.google.com/document/d/e/2PACX-1vQ6AOvKa_fExOKzgrr3Y05Ha4Neh0xi62rk-xpBirNA30Y2VrCRkLgL0fMn4_TiCW3_oo6fGBBd5rS2/pub |
| Mercer | Saint Henry Consolidated Local | https://www.sthenryschools.org/ |
| | | https://www.sthenryschools.org/athletics.html |
| Mercer | Fort Recovery Local | https://www.fortrecoveryschools.org/Default.aspx |
| | | https://fortrecoveryathletics.org/teams/2924369/boys/football/varsity/schedule |
| Mercer | Celina City | https://www.celinaschools.org/CelinaCitySchoolsPandemicInformation.aspx |
| | | https://www.arbiterlive.com/Teams/Schedule/5462234 |
| Mercer | Marion Local | https://www.marionlocal.org/ |
| | | https://marionlocalathletics.com/teams/2926076/boys/football/varsity/schedule |
| Van Wert | Delphos City | https://www.delphoscityschools.org/Content2/reopening |
| | | https://www.facebook.com/permalink.php?story_fbid=1575047562674408&id=183995885112923 |
| | | https://www.facebook.com/permalink.php?story_fbid=1584860938359737&id=183995885112923 |
| Van Wert | Crestview Local | https://www.crestviewknights.com/ReopeningOurSchool2020-2021.aspx |

| | | |
|---|---|---|
| | | https://www.crestviewknights.com/AthleticDirectorsMessage.aspx |
| Van Wert | Lincolnview Local | https://www.lincolnview.k12.oh.us/featured/7 |
| | | https://lincolnviewathletics.com/teams/3002008/girls/volleyball/varsity/schedule |
| Van Wert | Van Wert City | https://www.vwcs.net/News/reopening-vwcs-2020-2021 |
| | | https://www.vwcs.net/vwathletics/Content/basketball-boys |
